# SC Johnson Guardian™ spatial repellent shows one-year efficacy against wild pyrethroid-resistant *Anopheles arabiensis*, with similar blood-feeding inhibition efficacy to Mosquito Shield™ in a Tanzanian experimental hut trial

**DOI:** 10.1101/2024.11.21.24317755

**Authors:** Johnson Kyeba Swai, Watson Samuel Ntabaliba, Emmanuel Mbuba, Hassan Ahamad Ngoyani, Noely Otto Makungwa, Antony Pius Mseka, John Bradley, Madeleine Rose Chura, Thomas Michael Mascari, Sarah Jane Moore

## Abstract

Spatial repellents (SR) that passively emanate airborne concentrations of an active ingredient within a space disrupt mosquito behaviors to reduce human-vector contact. A clinical trial of SC Johnson’s Mosquito Shield™ (Mosquito Shield) has demonstrated 33% protective efficacy against malaria in Kenya. Mosquito Shield lasts for one month, but a longer duration product would be advantageous for deployment by malaria control programs. SC Johnson’s Guardian™ (Guardian), is designed to provide longer continuous protection from disease-transmitting mosquitoes. We conducted an experimental hut trial to evaluate the efficacy of Guardian over 12 months and compared it to Mosquito Shield over one month against wild pyrethroid resistant malaria vector mosquitoes, using entomological surrogates of clinical efficacy to assess the potential public health utility of Guardian. The primary endpoint was the number of blood-feeding *An. arabiensis*, while secondary endpoints were number landing proportion of *An. arabiensis* mortality and proportion blood-fed. Over 12 months of continuous tests, Guardian reduced numbers of *An. arabiensis* blood-feeding by 83% (95% Confidence Interval (79 – 86)) and landing by 65% (59 – 70). Guardian also induced 20% mortality (18 – 22). Guardian was found to be superior to Mosquito Shield in reducing the number of blood-fed *An. arabiensis*, and had similar proportions of blood-fed mosquitoes and 24-hours mortality. Results indicate that Guardian effectively reduces blood-feeding and landing of wild pyrethroid resistant malaria vectors for 12-months and shows superior protective efficacy to Mosquito Shield on the blood-feeding endpoint. Experimental hut studies are suitable for comparative evaluations of new spatial repellent products because they precisely estimate entomological endpoints elicited by spatial repellents known to significantly impact vectorial capacity and disease transmission.

**Key questions:** *What is already known on this topic?:*

- There is increasing evidence that spatial repellents have public health value. Randomized control trials have shown that SC Johnson Mosquito Shield™ (Mosquito Shield), a transfluthrin based spatial repellent, is effective in reducing malaria and dengue transmission. A WHO policy recommendation for the spatial repellent intervention class would follow a WHO-commissioned systematic review of the data from clinical trials.
- Mosquito Shield is effective for 30 days but malaria transmission lasts for 6 to 12 months in most endemic regions. Therefore, a longer lasting product required only one deployment per transmission season would be more operationally feasible, especially in remote areas of Africa where malaria burden is highest.

*What are the new findings?:*

- This is the first study to provide evidence of a spatial repellent product that can reduce the number of wild pyrethroid resistant *An. arabiensis* blood-feeding by 83% and landing by 65% while inducing a 20% delayed mortality at 24 hours for a duration of one-year.
- The study also demonstrated that Guardian was superior to Mosquito Shield (tested in RCTs) in reducing number of blood-fed *An. arabiensis*.

*Impact on vector control practice or policy:*

- Guardian meets the preferred product characteristics for a public health spatial repellent. It lasts a full year, and it does not require daily user interaction. This likely will result in higher adherence and coverage with lower operational cost for implementation compared to Mosquito Shield or other shorter-duration spatial repellents.
- Guardian is expected to have public health value by impacting disease transmission when used as a vector control tool against malaria because it showed superior performance when compared to Mosquito Shield, which has demonstrated malaria reduction in clinical trials.

## Introduction

In the last twenty years, the widespread deployment of insecticide treated nets (ITNs) and implementation of indoor residual spraying (IRS) have substantially reduced the global malaria burden; yet, progress has stalled, and the World Health Organization (WHO) has emphasized the need for additional control tools ^1^. The use of IRS is declining despite its proven effectiveness, primarily due to high implementation costs ^2^. ITN use has remained largely unchanged since 2015 with only 56% of young children and pregnant women sleeping under a net in 2022 ^1^. New malaria vector control tools must address the current gaps in protection, global public health funding constraints, insecticide resistance, climate change, and the expanding range of malaria vectors.

Several new intervention classes are in the pipeline including attractive targeted sugar baits, endectocides, gene drive, and spatial repellents ^3^. The spatial repellent intervention class currently is advancing toward a WHO policy recommendation ^3^ and guideline for implementation based on evidence generated on a one-month duration passive emanator product called SC Johnson Mosquito Shield™ (Mosquito Shield). Spatial repellents are devices that continuously disperse an active ingredient into the air, sustaining concentrations that impact several mosquito behaviors that are important in the malaria transmission cycle, including host detection, landing, blood-feeding, survival, and reproduction ^4^. Spatial repellent products in the public health space have evolved from consumer-facing products such as mosquito coils or electricity-powered liquid vaporizers, to passive emanator products ^5^ that work continuously over longer periods of time without the need for daily interaction with the end-user.

Several clinical trials have been conducted to demonstrate the impact against disease and thus public health value of Mosquito Shield in Indonesia ^6^ and Kenya ^7^. Additionally, implementation studies on Mosquito Shield in Rwanda and Syria, and on Guardian in Yemen, Nigeria, and Kenya, have evaluated effectiveness, coverage, acceptability, distribution strategies and cost, aiming to address knowledge gaps about their use in hard-to-reach displaced populations ^8^. Entomological efficacy trials conducted in the laboratory, semi-field, experimental hut ^9^, and in-home tests ^10^ have shown that Mosquito Shield provides substantial protection from mosquito bites throughout its one-month lifespan.

Whilst the public health value of spatial repellents in general, and Mosquito Shield specifically, as tools against malaria is becoming clearer, the WHO has identified longer lasting spatial repellents as a priority ^11^. Ideally a spatial repellent should be effective for 6 months or more, comparable to the duration of IRS ^5^. The SC Johnson Guardian™ (Guardian) was designed to provide continuous protection against malaria vectors bites for at least one full malaria transmission season (≥6-months), simplifying programmatic deployment. Should Mosquito Shield be determined by WHO as the first-in-class spatial repellent then Guardian must demonstrate that it performs better or no-worse than the first-in-class product on a primary end-point to join the spatial repellent class ^12^.

WHO requires a comparative analysis to assess the entomological performance of new products against a WHO prequalified comparator, using data from entomology studies already required for product prequalification ^12^. These entomological data provide indirect evidence of public health value using entomological surrogates of clinical efficacy and reassurance that a second-in-class product can offer a similar impact to the first-in-class product with proven epidemiological effects.

Here we report results from a 12-month evaluation of Guardian and compare its efficacy to a one-month evaluation of Mosquito Shield in experimental huts conducted in the same location against a population of pyrethroid-resistant *An. arabiensis* mosquitoes in Tanzania.

## Methods

### Study area

The study was conducted from May 2022 to May 2023 at Ifakara Health Institute’s Vector Control and Product Testing Unit (IHI-VCPTU) experimental hut site in Lupiro village (8.385°S, 36.670°E), Ulanga District, southeastern Tanzania. The local primary malaria vectors are *An. arabiensis* (>99.9% of the *An. gambiae* complex) and *An. funestus* (>80% *An. funestus* s.s.) ^9^. The dominant vector, *An. arabiensis* was resistant (<60% mortality) to alphacypermethrin, deltamethrin, lambda-cyhalothrin, and permethrin at 1x WHO discriminating doses at the time of trial conduct, but susceptibility was fully restored with pre-exposure to piperonyl butoxide (PBO) (S1 Table). Their resistance is attributed to CYP450 upregulation ^13^.

### Experimental huts

The study utilized the “New” Ifakara experimental huts (NIEH) which are the same design as the original Ifakara experimental huts ^14^ but divided with a fully sealed plywood wall to make two huts (Fig 1). The dimensions of the huts are 3.25 m x 3.5 m x 2 m (length x width x height) with a gabled roof of 0.5 m apex and volume of 28.43 m^3^. Each hut has 10 cm-wide eave gaps on three sides fitted with baffles that allow mosquitoes to enter freely, but they can only exit into two window traps allowing measurement of endpoints in the majority of mosquitoes that enter the huts ^15^. Both halves of a single original hut received identical treatments. Temperature and humidity were continuously monitored in one of the huts throughout the study.

**Figure 1.**
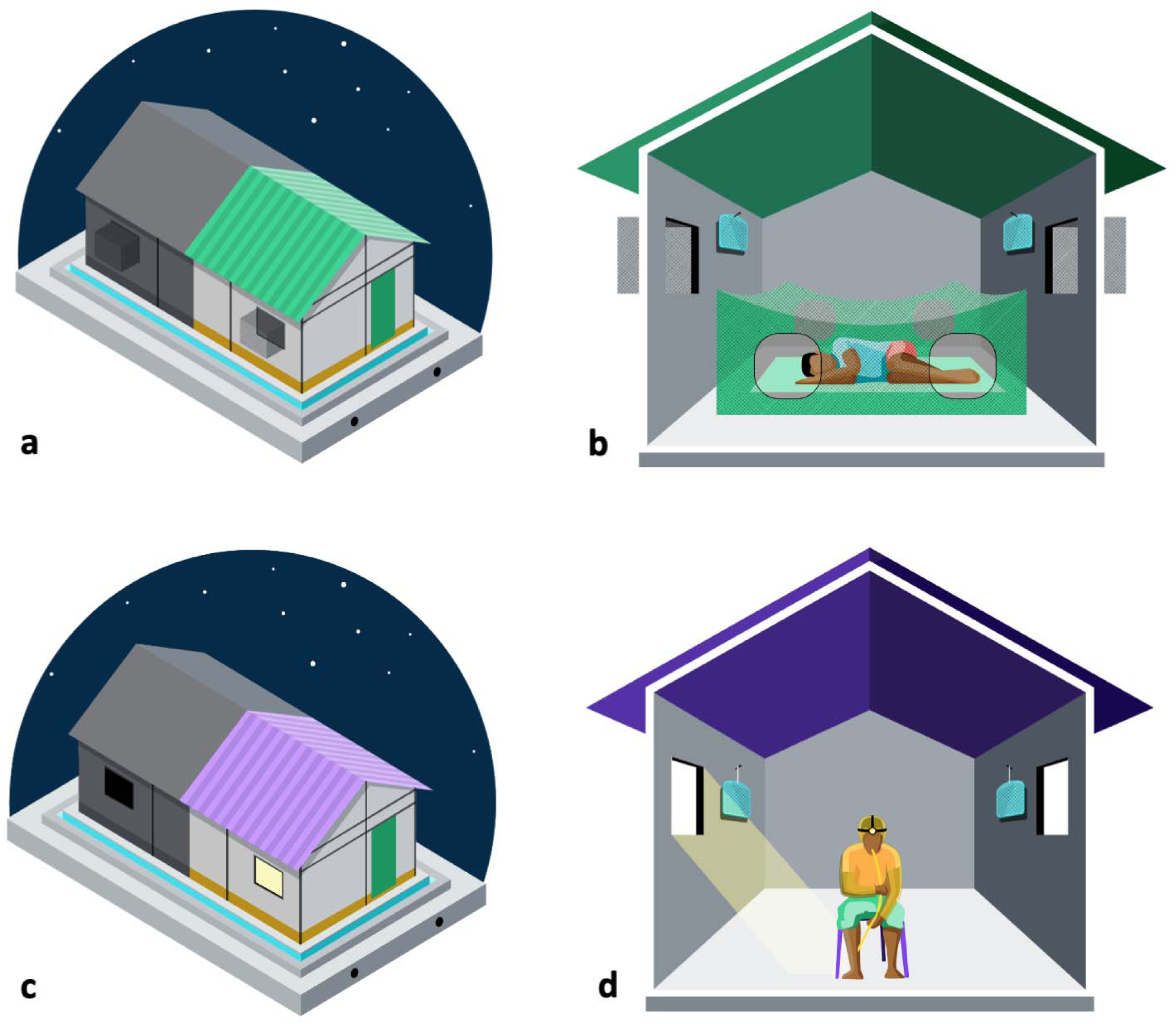
Set up of huts used for classic experimental hut “Feeding experiment” (a, b) and Human Landing Catch “Landing experiment” (c, d) including the placement of the Guardian™ (b, d)

### Intervention

Guardian is a passive emanator spatial repellent product that contains 2500 mg of transfluthrin on a mesh substrate within a plastic frame. Two Guardians were placed in each of the huts assigned to the treatment arm at a height of 1.5 m (Fig 1). The Guardian products were installed at 16:00 h on the 9th May, 2022 and were tested continuously through to 19th May, 2023.

Mosquito Shield is a passive emanator spatial repellent product dosed with 110 mg of transfluthrin in a folded 21.6 cm x 26.7 cm sheet of plastic film with a label claim of one month. Four Mosquito Shield products were placed according to manufacturer specifications at a height of 1.8 m from the ground and at center length of each wall in each hut: one on each of the walls. The Mosquito Shield products were installed at 16:00 h on the 26 November, 2022 and were removed after 32 days.

Both spatial repellent products are manufactured by SC Johnson & Son, Racine, WI, USA.

### Study design

#### Guardian

The study employed a partially randomized block time-series design with two treatment groups (Guardian and no product as control) across eight huts per arm (16 huts in total). The huts were divided into four sets of four huts (two treatment, two control), within which volunteers rotated every night in a 4×4 Latin square design to address varying mosquito attractiveness between individuals. Treatments were assigned to huts sequentially to minimize spatial bias and remained in the same huts throughout the study. The primary endpoint was number of blood fed mosquitoes; secondary endpoints were proportion of mosquitoes blood feeding and the proportion of mosquitoes dead at 24 hours post exposure. In addition, the number of mosquitoes landing was measured in a parallel experiment as human landing can be measured operationally, whereas mosquito blood feeding and mortality cannot ^16^. The efficacy of Guardian was evaluated for 12-months, using two methods in the same huts: 1) a feeding experiment measuring mosquito blood feeding and mortality, and 2) a landing experiment measuring mosquito landings on volunteers. At the start of every month, the feeding experiment was run first followed by the landing experiment, each lasting eight consecutive nights.

### Feeding experiment

Each test night 16 male volunteers slept in huts from 1800-0600 h under untreated, deliberately-holed “too-torn” bed nets (6 holes of 25 by 25 cm, making >707 cm^2^ surface area) (Fig 1 a & b). Mosquitoes inside the bed net or in window traps were collected in the morning using mouth aspirators and those resting on the walls or knocked down on the floor using Prokopack aspirators. These were then sorted by the location that they were collected from and by physiological status (dead unfed, dead fed, alive unfed, or alive fed); then held at a 24.7 (23.7 – 25.4) °C (median (IQR)) with access to 10% sucrose solution for 24 hours. *Anopheles* species females were morphologically identified to species level ^17^. A subsample of 105 mosquitoes was submitted to the laboratory for speciation using polymerase chain reaction (PCR) ^18^.

### Landing experiment

On each test night, 32 male volunteers conducted Human Landing Catches (HLC) in the 16 huts (Fig 1 c & d). Volunteers worked in shift-pairs, performing HLC for six hours every night i.e., from either 1800 - 0000 h or 0000 - 0600 h. Volunteers sat at the center of each hut exposing only their lower legs while wearing closed-toe shoes and net jackets to standardize the area that mosquitoes could land on. Using mouth aspirators and torches, volunteers captured mosquitoes landing on their legs for 50 min of each hour, and took a 10-min refreshment break. Each hour, the captured mosquitoes were taken to the field laboratory and incapacitated in a freezer. The following morning *Anopheles* species were morphologically identified to species level ^17^ and counted. The huts had no window exit-traps and the windows were left open at night to maximize mosquito entry. During the day the windows were kept closed between 0700 h and 1600 h, leaving eaves open for airflow like local homes.

#### Mosquito Shield

An independent experimental hut test of Mosquito Shield was run for its full duration of efficacy (32 days) mid-way through the Guardian evaluation, between November and December 2022 (Supplemental Fig 1). Mosquito collection was done as described in the feeding experiment above. Data collected from this and the Guardian feeding experiment above were used for the comparison of Guardian to Mosquito Shield.

**Supplemental Figure 1** Study timelines for the two independent contemporaneous experimental hut studies used for the comparison of Guardian versus Mosquito Shield.

Mosquito Shield was evaluated in a total of 8 huts. Half of the huts (4) received no intervention (control) while the remaining (4 huts) received the intervention (four Mosquito Shield). The treatments (Mosquito Shield, or untreated control) were randomly allocated to huts using a random number generator and remained fixed in the huts for the duration of the study. The primary endpoint was number of blood fed mosquitoes, and secondary endpoints were proportion of mosquitoes blood feeding and the proportion of mosquitoes dead at 24 hours post exposure. A total of eight study participants rotated sequentially through the eight huts (4 control and 4 treatment).

### Sample size

Sample size calculations were performed using simulation-based power analysis in R statistical software with significance level of 0.05 for rejecting the null hypothesis. For the evaluation, 1,000 simulations for generalized linear mixed models [6] were run using a Latin square design with volunteers rotating nightly. Variances were set at 0.14 for hut, 0.61 for daily observation and 0.21 variation in attraction to mosquitoes among volunteers, based on previous observations and an estimated 16 *An. arabiensis* mosquitoes caught per night. The study was powered to detect a 30% difference in mosquito blood-feeding between the intervention arm and negative control each month assuming 60% feeding rate in the control.

The Guardian study was run for 8 huts per arm for 8 nights per month to give 64 hut nights per month per treatment arm. The Mosquito Shield experiment was run continuously for 32 nights in 4 huts per treatment arm to give 128 hut nights per treatment arm. Simulations indicated that the independent Guardian and Mosquito Shield studies were powered at 100% (95% CI: 100 - 100).

Post hoc power analysis with 1,000 simulations for generalized linear mixed models [6] were run using a Latin square design for each experiment. The following estimates observed from the Guardian study were used for the analysis: study variation of log of 1.034, mosquito distribution estimate of log of 0.92, a geometric mean of 15 *An. arabiensis* mosquitoes caught per night, 13% blood-fed mosquitoes in intervention arm and 29% blood-fed mosquitoes in the negative control arm. The following estimates observed from the Mosquito Shield study were used for the analysis: study variation of log of 0.9905, mosquito distribution estimate of log of 0.53, a geometric mean of 7 *An. arabiensis* mosquitoes caught per night, 15% blood-fed mosquitoes in intervention arm and 34% blood-fed mosquitoes in the negative control arm. Post hoc power of the Guardian study was 100% (95% CI: 99-100) and Mosquito Shield experiment was 98% (95% CI: 97 – 99).

### Analysis

All analysis was performed using STATA^®^ 18 software (StataCorp LLC, USA). We analyzed the Guardian longitudinal entomological efficacy data as follows: data distribution was checked using histograms and measures of variance relative to the mean. Williams means ^19^ with 95% confidence intervals (95% CI) were calculated for numbers of mosquito landings and blood-feeding. For 24-hour mortality and blood feeding proportions, arithmetic means with 95% CI were estimated. Control corrected 24-hour mortality was not estimated since the mortality in the control arm was <5%. Regression analysis used a mixed effect negative binomial regression model for count outcomes and logistic regression models for proportion outcomes. The models included treatment, volunteer, and night as fixed effects, and hut as a random effect to account for clustering at the level of hut because interventions were fixed for the duration of the study. Protective efficacy was assessed by computing blood-feeding inhibition and landing inhibition using the formula (1 – IRR/OR) x 100, where IRR & OR respectively represent incidence rate ratios and odds ratio in the Guardian arm compared to the control arm.

For the comparison of Guardian to Mosquito Shield, we chose protective efficacy estimated in terms of the reduction in the number of blood-fed mosquitoes captured in the experimental hut as our primary outcome ^15^. This is because spatial repellents reduce mosquito house entry as well as the ability of mosquitoes to blood-feed when they are inside a space ^4^ ^20^. Secondary outcomes were protective efficacy in terms of reduction in the proportion of mosquitoes blood-fed and increased proportion of mosquitoes dead at 24 hours post collection from the experimental huts, similar to those measured for ITNs ^21^. We examined the effect of Guardian and Mosquito Shield on the number of mosquitoes blood-fed using a mixed effects negative binomial regression. For the secondary outcomes which were proportions, mixed effects logistic regression was used. Intervention, volunteer, and experimental night were included as fixed categorical factors. Hut was added as a random factor to account for clustering of observations because interventions were fixed for the duration of the study and a dummy variable created to distinguish the Guardian or Mosquito Shield treatments was interacted with treatment variable.

### Results

Confirmatory sub-species identification using PCR showed that 100% (102/102) of the amplified subsample of *An. gambiae* s.l were *An. arabiensis*.

In the Guardian feeding experiment a total of 26,920 *An. arabiensis* mosquitoes were caught in the control arm and 12,863 in the Guardian arm. In the Guardian landing experiment, a total of 67,857 *An. arabiensis* mosquitoes were caught in the control arm and 29,724 in the Guardian arm. In the Mosquito Shield feeding experiment, a total of 4,557 *An. arabiensis* mosquitoes were caught in the control arm and 3,205 in the Mosquito Shield arm.

### Protective Efficacy of Guardian

We observed that over 12-months, Guardian provided 82.7% 95% CI (78.5 – 86.1%) protection from mosquito bites and 65.1% (59.4 – 70.0%) reduction in the number of mosquitoes landing on human volunteers (Table 1). The blood-feeding and landing protective efficacies of Guardian were statistically significant for the combined 12-months data (Table 1) and every individual month (Fig 2, Supplemental Table 2 & Supplemental Table 3). Each month, the protective efficacy measured by reduction in number blood feeding was higher than that measured by reduction in number landing (Figure 2). Guardian killed 20.1% (18.4 – 21.8%) of the *An. arabiensis* in the study area even though this mosquito population is strongly resistant to pyrethroid insecticides. In addition, only 12.7% (11.4 – 14.1%) of mosquitoes that entered huts with Guardian blood-fed over the 12-months trial (Table 2). Statistically significant higher mortality and lower blood-feeding proportions were also observed in the Guardian arm overall, and during each month of the evaluation (Supplemental Table 4 & Supplemental Table 5).

**Figure 2.**
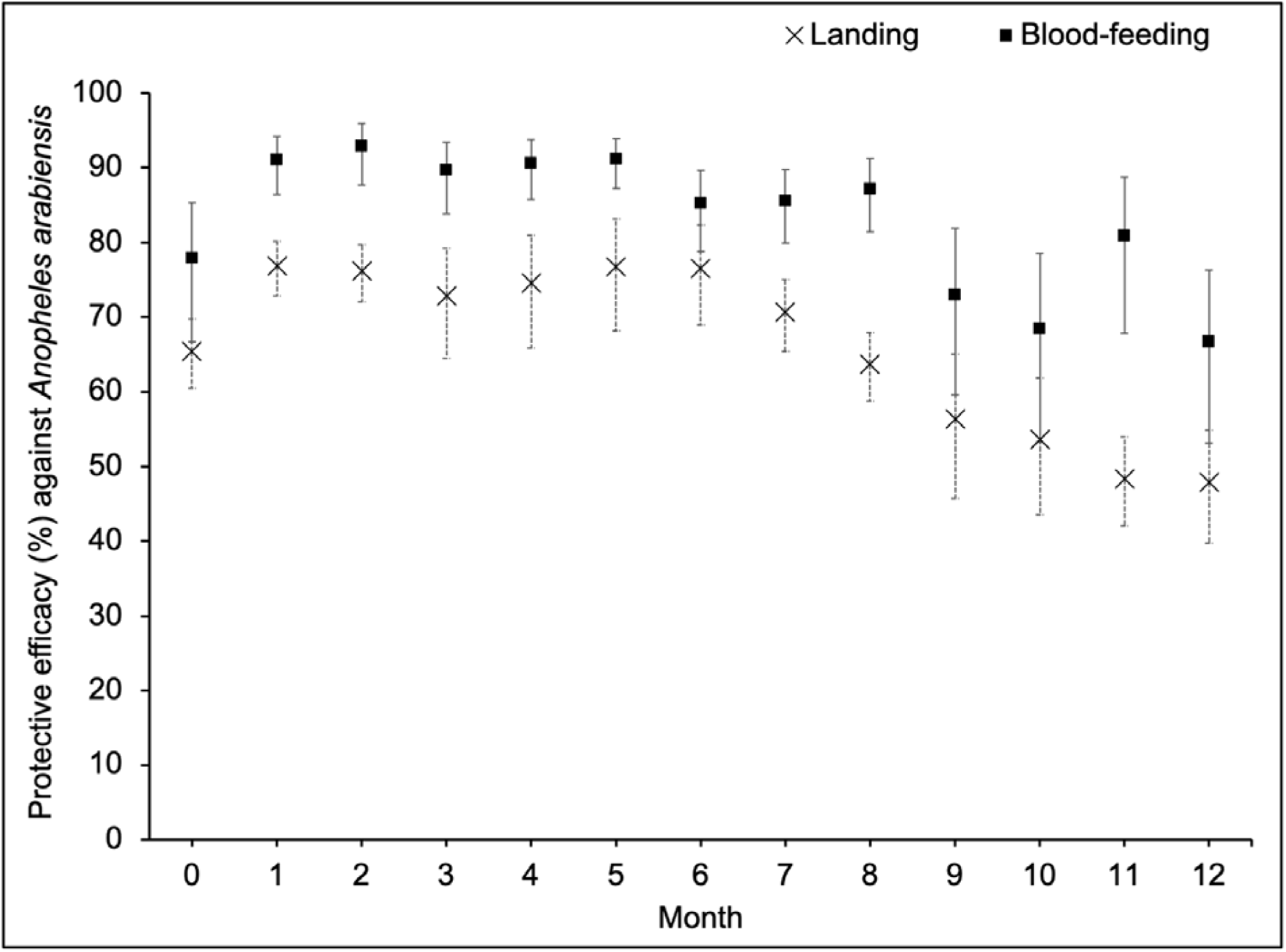
Monthly trend of landing and blood-feeding inhibition protective efficacy of Guardian against wild pyrethroid-resistant *An. arabiensis*.

**Table 1.**
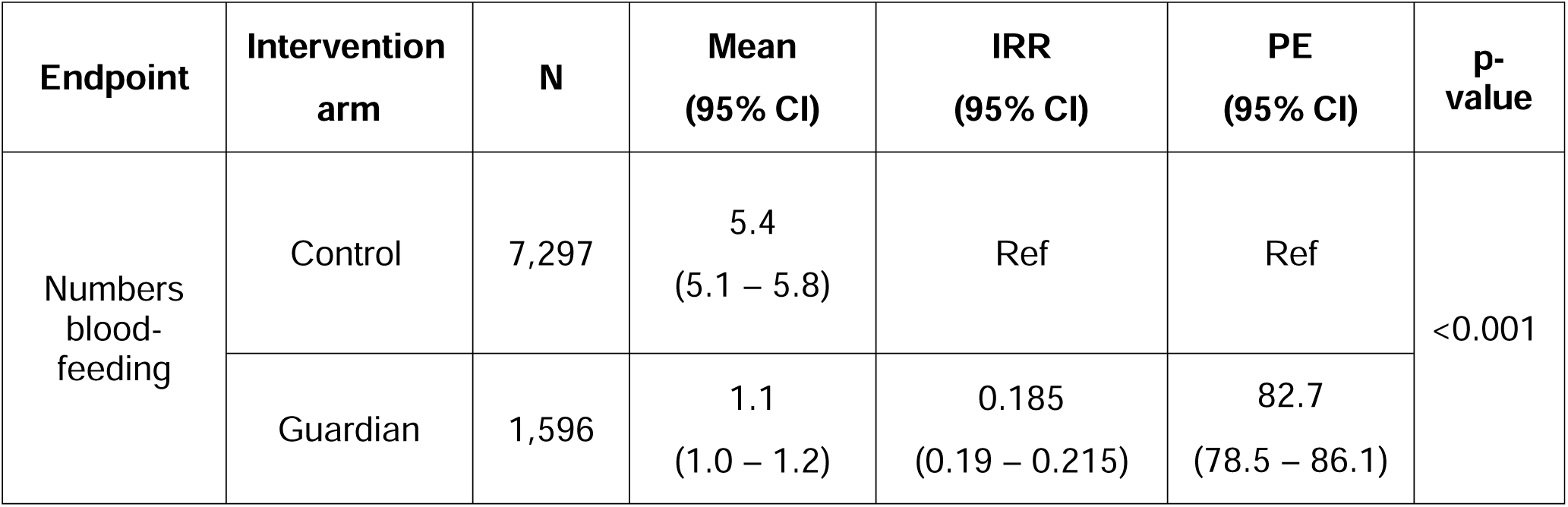
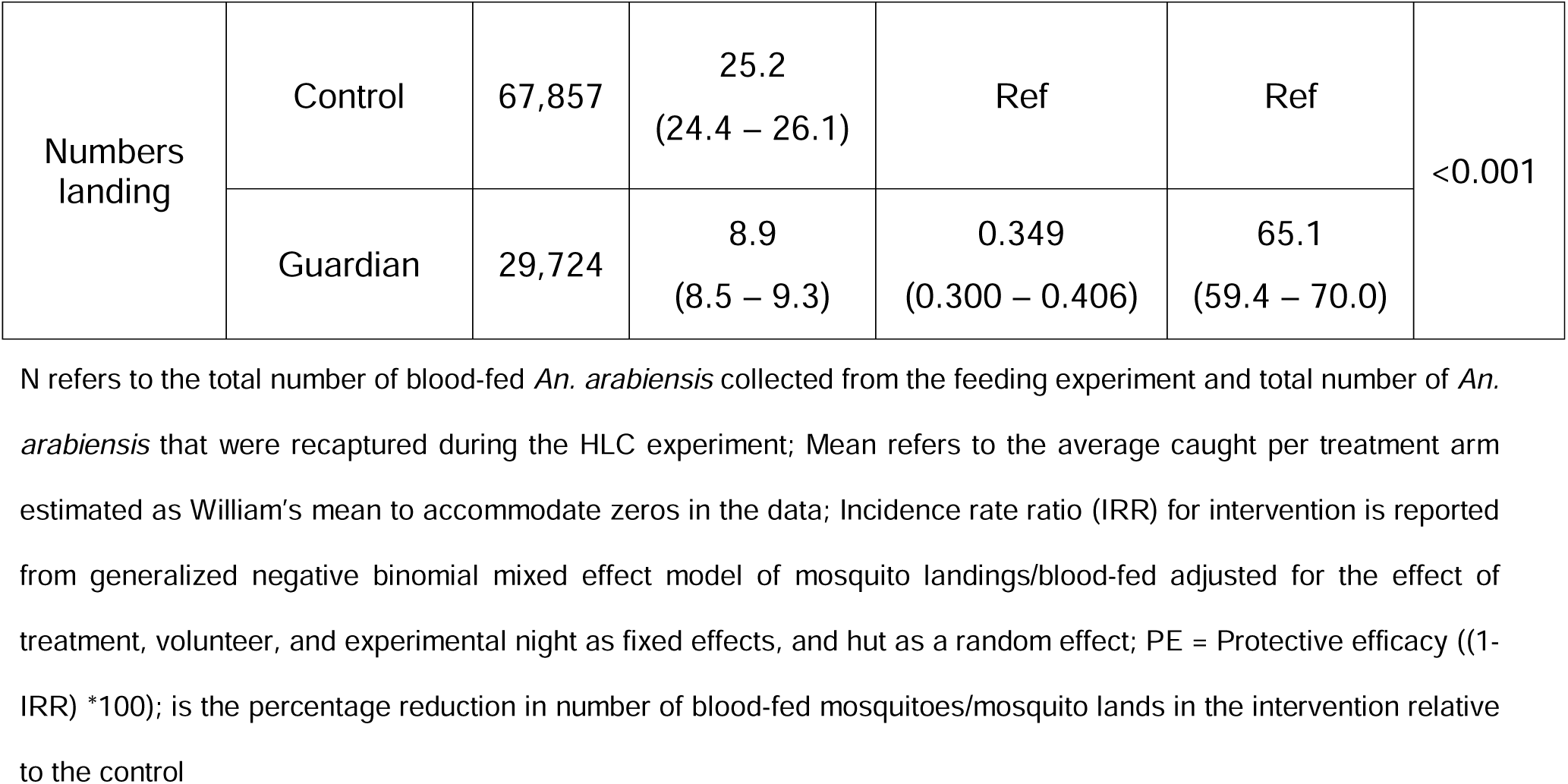
Protective efficacy of Guardian™ in reducing blood-feeding and human landings of wild pyrethroid-resistant *An. arabiensis* over 12 months.

**Table 2.**
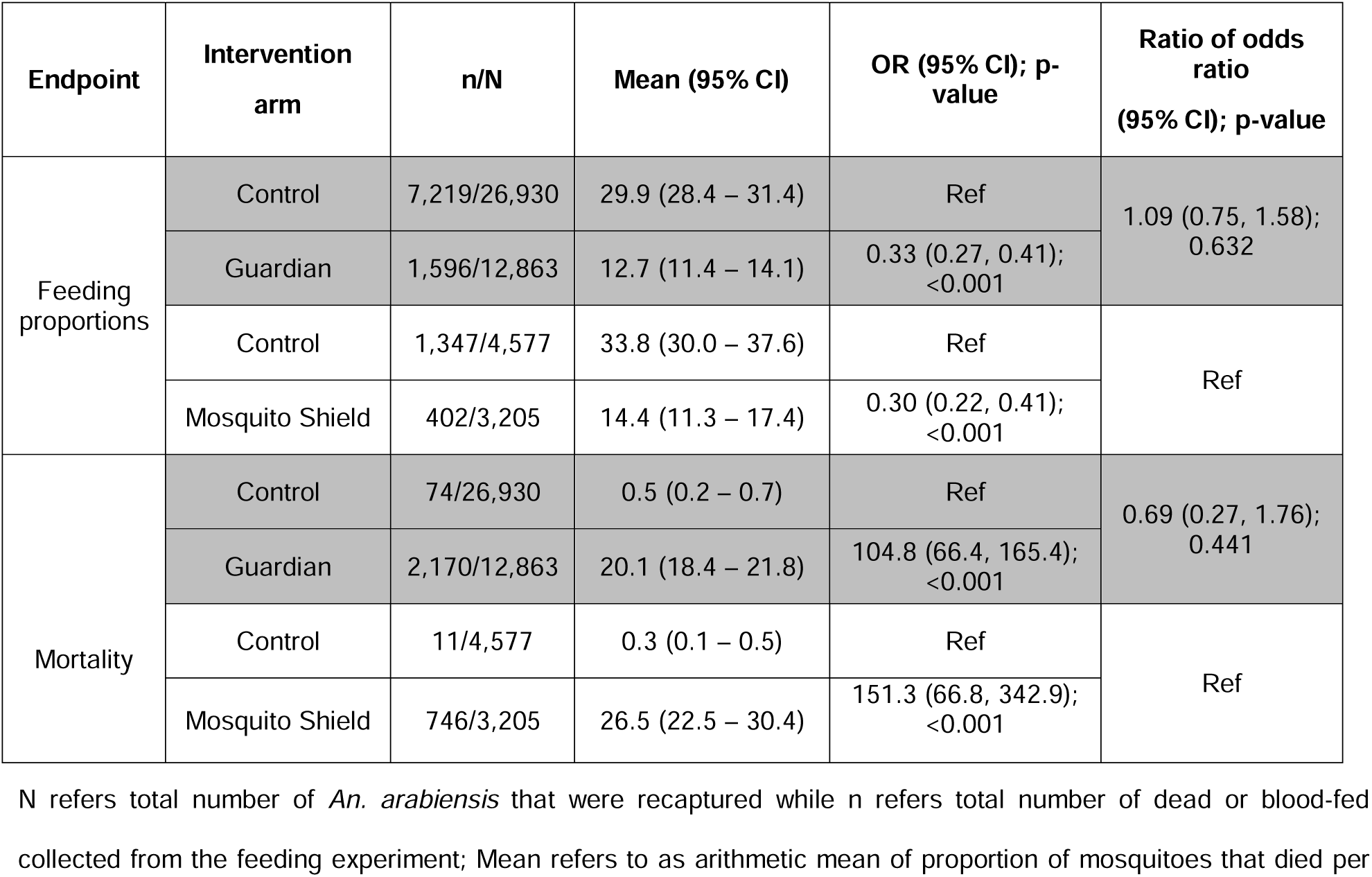
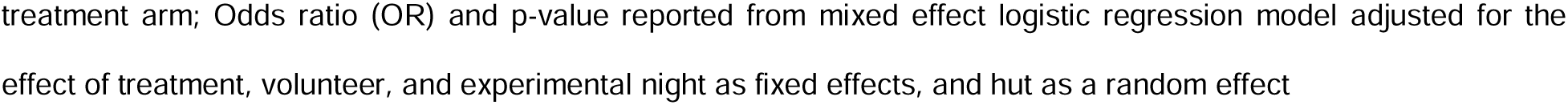
Guardian induced mortality at 24 hours and proportion blood fed (in percentage) of wild pyrethroid-resistant *An. arabiensis* and its comparison relative to Mosquito Shield™.

### Comparison of Guardian to Mosquito Shield

For the primary endpoint of the rate of blood-fed mosquitoes, the effect of Guardian (0.17 (0.14, 0.21), p<0.001) was larger than the effect of Mosquito Shield (0.29 (0.21, 0.41), p<0.001) (Table 3). The ratio of the rate ratios was 0.60 (0.40, 0.89), p=0.012 (Table 3), showing evidence that Guardian is superior in reducing the number of blood-fed mosquitoes.

**Table 3.**
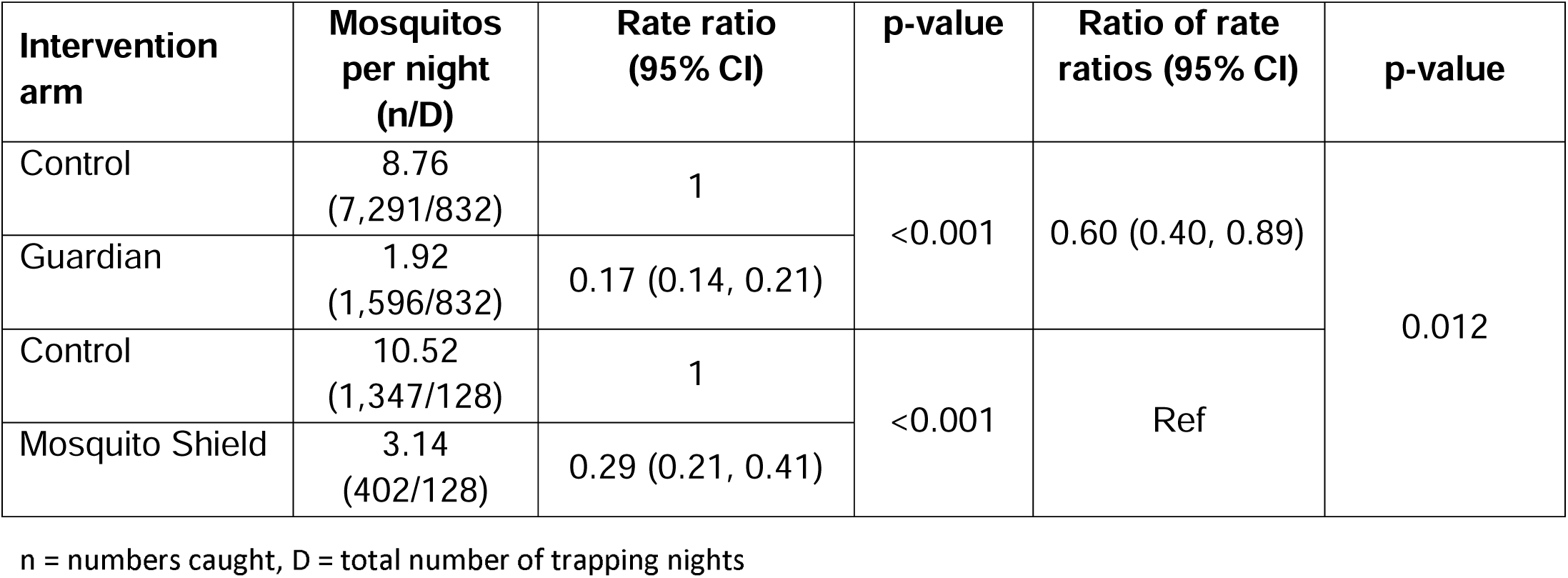
Comparison of Guardian™ relative to Mosquito Shield™ over the duration of life in reducing number of wild pyrethroid-resistant *An. arabiensis* blood-feeding.

For the secondary endpoints, there was no evidence Guardian performed differently to Mosquito Shield. The ratio of the odds for proportion of blood-fed, was 1.09 (0.75, 1.58), p=0.632 and for induced mortality was 0.69 (0.27, 1.76) p=0.441 (Table 2).

## Discussion

### Summary of the study

In this study we evaluated the efficacy of the spatial repellent Guardian against a pyrethroid-resistant population of *An. arabiensis* in experimental huts in Tanzania over 12-months. We additionally compared the efficacy of Guardian to Mosquito Shield that has demonstrated public health benefit in randomized control trials ^3^. Experimental huts simulate residential settings, allowing us to evaluate vector control tools under standardized conditions in which wild free-flying mosquitoes enter a human habitation and interact with a human in the presence of an intervention. They also enable the direct measurement of several endpoints elicited in mosquitoes by spatial repellents, including reduced blood-feeding and induced mortality which significantly impact vectorial capacity ^22^ and are linked to epidemiological outcomes ^23^. These outcomes are more difficult to measure operationally, unless population level effects are accurately measured ^24^.

### Duration of effect and its relevance to public health

The study found that Guardian substantially reduced the number of blood-feeding (83%) and landing (65%) pyrethroid-resistant malaria vectors for up to one-year, consistent with the preferred product characteristics for public health use ^5^. Longer-lasting products require less frequent replacement, reducing operational costs and increasing the likelihood of sustained protection. Consistent use by households is more likely for interventions that offer long term efficacy with no daily behavioral or maintenance requirements. Indeed, the challenge of sustaining high levels of consistent use is the major reason that topical repellents have not shown public health benefit ^25^. The one-year duration of Guardian exceeds the observed duration for some IRS formulations currently available ^26^, although it is shorter than the lifespan of most ITNs ^27^. In addition, once placed it will provide round the clock protection against even day biting malaria transmitting mosquitoes ^28^ ^29^, a gap in protection conferred by ITN and IRS which mostly target night biting mosquitoes. Guardian can be installed by community health workers or household members without technical training or personal protective equipment, and it can be deployed using similar channels to those currently used for ITNs.

### Endpoints measured

Throughout the study, blood-feeding inhibition was consistently higher than landing inhibition. This finding has also been observed in other studies of transfluthrin-based hessian emanators ^30 31^ and in a previous study of Mosquito Shield ^15^. Pyrethroids also reduce mosquitoes’ blood feeding in the presence of human hosts under damaged bed nets ^32^, or near pyrethroid treated nets ^33^, by affecting olfactory neurons ^34^. Ignoring secondary impacts of pyrethroid based spatial repellents likely underestimates their impact on vectorial capacity ^31^. While HLC does expose humans to risk of disease vectors, it is currently the only feasible means of operationally evaluating the impact of spatial emanators on mosquito populations, as light traps are ineffective for this purpose ^16^.

### Resistance

In the study area, like much of sub-Saharan Africa, pyrethroid-resistance is present. Even so, Guardian killed 20% of *An. arabiensis* mosquitoes over the duration of 12-months, similar to that of pyrethroid bed nets tested in this location ^35^. Transfluthrin, the active ingredient in Guardian, is structurally divergent from the majority of pyrethroids as it has a polyfluorobenzyl moiety and remains active in mosquitoes with upregulated P450 levels that can metabolically detoxify most pyrethroids used in public health ^36^ and has been found to be resilient to mechanisms that drive resistance in *An. funestus* ^37^: a highly efficient malaria vector known to be resistant to other pyrethroids used in public health ^38^. This supports the use of transfluthrin-based products in areas of known pyrethroid-resistance, although mosquito mortality will be dependent on both dose and mosquito resistance profile ^31^.

### Policy

Comparative evaluations of second-in-class ITNs and IRS to their respective first-in-class for which there is epidemiological evidence of public health benefit are carried out in experimental hut trials ^12^. We compared Guardian’s efficacy to Mosquito Shield, finding Guardian superior in reducing successive blood-feeding in *An. arabiensis*. If WHO develops a guideline for use of spatial repellents as an intervention class for use in malaria control, our data suggest Guardian would also have a significant epidemiological impact. The study adapted ITN experimental hut methods to evaluate indoor passive spatial repellents like Guardian, considering hut as a random effect due to treatments remaining fixed in the same huts.

### Limitations and research gaps

The scope of this study was limited to measuring the entomological efficacy of Guardian on *An. arabiensis* mosquitoes in Tanzania. Future work with Guardian should consider measuring the impact against other malaria vectors in different geographical areas and in different contexts of insecticide resistance.

Evaluation of Guardian’s operational effectiveness when deployed alone or in combination with core malaria control interventions (ITNs and IRS) as part of integrated vector management will be critical as it is likely to have an additive effect when combined with other interventions ^39^. Monitoring continued effectiveness post-deployment, especially through developing cost-effective chemical and laboratory assays analogous to those used for operational monitoring of ITNs ^40^ is a research priority. In addition, mathematical modeling of the impact of spatial repellents in different epidemiological contexts with different intervention mixes could help inform national strategic plans and sub-national tailoring by country malaria control programs. Further operational research is ongoing to evaluate the impact that Guardian may have for people at risk of malaria, dengue and leishmaniasis that are in need of humanitarian assistance and living in temporary shelters ^41^. Additional information from ongoing and future deployments, as well as operational research on Guardian will better inform distribution strategies, cost of implementation, and operational effectiveness against vector borne disease.

## Conclusion

This study demonstrated that Guardian was efficacious in reducing number of blood-fed wild pyrethroid-resistant *An. arabiensis* mosquitoes in Tanzania for up to one-year. This study supports transfluthrin-based spatial repellents as additional vector control tools, offering easily transported, compliance-free protection. This is a major advancement in the field of public health as low compliance is the main reason that repellents have previously not shown disease reduction in trials. Comparative efficacy data suggest that Guardian can be expected to provide similar public health benefit to Mosquito Shield, which has been demonstrated to reduce malaria transmission in East Africa ^42^ and *Aedes*-borne virus transmission in Peru ^43^. The study also outlines a methodology to measure the efficacy of similar products comparable to that used for the evaluation of ITNs and IRS that precisely estimates the primary endpoint of blood-feeding reduction as well as mortality that both substantially impact the vectorial capacity and ultimately disease transmission by mosquito vectors.

## Data Availability

All data produced in the present study are available upon reasonable request to the authors

## Ethics statement

Written informed consent was obtained from all study participants prior to commencement of the study. All study volunteers were provided with Doxycycline® malaria prophylaxis as per Tanzania Guidelines for Diagnosis and Treatment of Malaria and tested weekly for malaria infection using malaria rapid diagnostic tests (SD Bioline) administered by a medical officer. No volunteers tested positive for malaria throughout the duration of this study. No adverse effects were reported among the volunteers throughout the duration of the study. The study did not involve patients or the public in its design, conduct, reporting or dissemination plans. Study activities were approved by the Institutional Review Board of IHI IHI/IRB/EXT/No: 34 – 2022 and National Institute for Medical Research Tanzania NIMR/HQ/R.8c/VOL.I/2177.

## Availability of data and materials

The datasets used and or analyzed in this study are available from the corresponding authors upon reasonable request.

## Acknowledgement

The study was supported by S. C. Johnson & Son, Inc, Racine, Wisconsin. The authors express their sincere thanks and appreciation to the study volunteers, who worked tirelessly over the duration of the experiment. Village leaders and community which surrounds the Ifakara Health Institute experimental hut site in Lupiro, for allowing us to continually run our experiments with minimal interruptions. A special thanks to the Vector Control and Product Testing Unit (VCPTU) management, administrators and colleagues who helped in organizing logistics and materials allowing smooth performance of the study.

## Author contribution

Designed study: JKS, and SJM. Conducted the study: JKS, WSN, HAN, NOM, and APM, Statistical analysis: JKS, JB & SJM. Wrote manuscript: JKS, & SJM. Critically reviewed final draft: JB, MRC, TMM & SJM. EM also oversaw that all project activities were carried out following Good Laboratory Practice (GLP). All authors read and approved the final manuscript.

## Consent for publication

Permission to publish this study was obtained from National Institute for Medical Research Ref No BD.242/437/01C/33 under ethical clearance NIMR/HQ/R.8c/VOL.I/2177.

