## Supplemental Table 1 for "SC Johnson Guardian™ spatial repellent shows one-year efficacy against wild pyrethroid-resistant *Anopheles arabiensis*, with similar blood-feeding inhibition efficacy to Mosquito Shield™ in a Tanzanian experimental hut trial": Supplemental Table 1 .docx

**S1 Table** WHO susceptibility test results for *An. arabiensis* at the Lupiro experimental hut at the beginning (2022) and end (2023) of the study.

| Year | Outcome | Permethrin (0.75%) | Deltamethrin (0.05%) | $\alpha$-cypermethrin (0.05%) | $\lambda$-cyhalothrin (0.05%) | Pirimiphos-methyl 170mg/m^2^ | Bendiocarb (0.1%) |
| --- | --- | --- | --- | --- | --- | --- | --- |
| 2022 | KD60 | 79% | 15% | 14% | 61% | 45% | 94% |
|  | M24 | 51% | 18% | 5% | 45% | 100% | 100% |
| 2023 | KD60 | 66% | 55% | 54% | 5% | 17% | 100% |
|  | M24 | 13% | 18% | 5% | 8% | 100% | 100% |
