## Supplemental Table 2 for "SC Johnson Guardian™ spatial repellent shows one-year efficacy against wild pyrethroid-resistant *Anopheles arabiensis*, with similar blood-feeding inhibition efficacy to Mosquito Shield™ in a Tanzanian experimental hut trial": Supplemental Table 2 .docx

**S2 Table** Monthly protective efficacy of Guardian in reducing number of wild pyrethroid-resistant *An. arabiensis* landing.

| Month | Intervention | Total captured | Mean (95% CI) | % PE (95% CI) | p-value |
| --- | --- | --- | --- | --- | --- |
| 0 | Control | 2,614 | 16.6 (14.8 - 18.7) | _ | <0.001 |
|  | Mesh | 929 | 5.2 (4.3 - 6.2) | 65.4 (60.5, 69.7) |  |
| 1 | Control | 2,599 | 16.9 (15.0 - 19.0) |  | 0.001 |
|  | Mesh | 625 | 3.5 (2.9 - 4.1) | 76.8 (72.8, 80.2) |  |
| 2 | Control | 3,238 | 20.0 (17.6 - 22.8) |  | <0.001 |
|  | Mesh | 840 | 4.6 (4.0 - 5.5) | 76.1 (72.0, 79.7) |  |
| 3 | Control | 3,690 | 24.6 (22.2 - 27.3) |  | <0.001 |
|  | Mesh | 1,094 | 6.2 (5.2 - 7.2) | 72.8 (64.5, 79.2) |  |
| 4 | Control | 5,539 | 37.0 (33.4 - 41.1) |  | <0.001 |
|  | Mesh | 1,591 | 9.0 (7.7 - 10.5) | 74.5 (65.9, 81.0) |  |
| 5 | Control | 3,596 | 23.8 (21.3 - 27.6) |  | <0.001 |
|  | Mesh | 911 | 5.2 (4.5 - 6.1) | 76.7 (68.1, 83.1) |  |
| 6 | Control | 1,990 | 13.1 (11.8 - 14.5) |  | <0.001 |
|  | Mesh | 475 | 2.8 (3.4 - 4.3) | 76.5 (68.9, 82.3) |  |
| 7 | Control | 3,409 | 20.6 (18.0 - 23.4) |  | <0.001 |
|  | Mesh | 1,052 | 5.8 (4.9 - 6.8) | 70.6 (65.4, 75.0) |  |
| 8 | Control | 7,242 | 47.7 (42.8 – 53.1) |  | <0.001 |
|  | Mesh | 2,861 | 17.0 (14.9 – 19.5) | 63.7 (58.8, 67.9) |  |
| 9 | Control | 6,384 | 41.4 (37.0 – 46.4) |  | <0.001 |
|  | Mesh | 3,228 | 17.5 (15.0 – 20.5) | 56.4 (45.7, 65.0) |  |
| 10 | Control | 6,204 | 42.4 (38.4 – 46.7) |  | <0.001 |
|  | Mesh | 3,332 | 19.0 (16.4 – 22.0) | 53.6 (43.6, 61.8) |  |
| 11 | Control | 6,067 | 36.5 (31.8 – 41.9) |  | <0.001 |
|  | Mesh | 3,347 | 18.5 (15.8 – 22.6) | 48.4 (42.1, 54.0) |  |
| 12 | Control | 4,209 | 25.4 (22.2 – 29.1) |  | <0.001 |
|  | Mesh | 2,231 | 13.1 (11.4 – 15.1) | 47.9 (39.8, 54.9) |  |
| Overall | Control | 56,786 | 26.1 (25.2 – 27.1) | _ | <0.001 |
|  | Mesh | 22,516 | 8.2 (7.8 – 8.6) | 65.1 (59.4, 70.0) |  |

Total caught = overall sum of mosquitoes catches via hourly HLC for each month and combined for the months; Mean= Average caught per night per hut estimated as Williams mean due to skewness of mosquito count data; PE = Protective efficacy (1-IRR) – it measures percentage reduction in mosquito landing on a participant in the intervention relative to the control.
