## Supplemental Table 3 for "SC Johnson Guardian™ spatial repellent shows one-year efficacy against wild pyrethroid-resistant *Anopheles arabiensis*, with similar blood-feeding inhibition efficacy to Mosquito Shield™ in a Tanzanian experimental hut trial": Supplemental Table 3 .docx

**S3 Table** Monthly protective efficacy of Guardian in reducing number of wild pyrethroid-resistant *An. arabiensis* blood-feeding.

| Month | Intervention | Total Fed | Mean (95% CI) | % PE (95% CI) | p-value |
| --- | --- | --- | --- | --- | --- |
| 0 | Control | 186 | 1.5 (1.0 – 2.1) | _ | <0.0001 |
|  | Mesh | 42 | 0.4 (0.2 - 0.6) | 77.9 (66.7, 85.3) |  |
| 1 | Control | 388 | 4.0 (3.0 - 5.2) |  | <0.0001 |
|  | Mesh | 36 | 0.4 (0.2 - 0.5) | 91.1 (86.4, 94.2) |  |
| 2 | Control | 328 | 2.8 (2.0 – 3.9) |  | <0.0001 |
|  | Mesh | 29 | 0.2 (0.1 - 0.4) | 92.9 (87.7, 95.9) |  |
| 3 | Control | 280 | 2.9 (2.2 – 3.8) |  | <0.0001 |
|  | Mesh | 29 | 0.3 (0.2 - 0.5) | 89.7 (83.8, 93.4) |  |
| 4 | Control | 830 | 9.0 (7.2 - 11.2) |  | <0.0001 |
|  | Mesh | 78 | 0.8 (0.5 - 1.1) | 90.6 (85.8, 93.8) |  |
| 5 | Control | 562 | 6.9 (5.7 - 8.4) |  | <0.0001 |
|  | Mesh | 51 | 0.5 (0.3 - 0.7) | 91.2 (87.3, 93.9) |  |
| 6 | Control | 331 | 4.3 (3.6 - 5.1) |  | <0.0001 |
|  | Mesh | 49 | 0.5 (0.3 - 0.7) | 85.2 (78.8, 89.7) |  |
| 7 | Control | 432 | 5.3 (4.3 - 6.4) |  | <0.0001 |
|  | Mesh | 63 | 0.7 (0.4 – 0.9) | 85.6 (79.9, 89.8) |  |
| 8 | Control | 677 | 8.0 (6.5 – 9.9) |  | <0.0001 |
|  | Mesh | 91 | 1.0 (0.7 – 1.3) | 87.2 (81.4, 91.2) |  |
| 9 | Control | 1,066 | 11.4 (8.8 – 14.8) |  | <0.0001 |
|  | Mesh | 358 | 3.1 (2.1 – 4.2) | 73.0 (59.6, 81.9) |  |
| 10 | Control | 812 | 10.0 (8.4 – 12.0) |  | <0.0001 |
|  | Mesh | 299 | 2.9 (2.1 – 3.9) | 68.4 (53.3, 78.6) |  |
| 11 | Control | 1,002 | 12.1 (10.0 – 14.7) |  | <0.0001 |
|  | Mesh | 335 | 2.5 (1.7 – 3.6) | 80.9 (67.8, 88.7) |  |
| 12 | Control | 403 | 4.0 (2.9 – 5.3) |  | <0.0001 |
|  | Mesh | 136 | 1.3 (0.9 – 1.8) | 66.7 (53.2, 76.3) |  |
| Overall | Control | 7,297 | 5.5 (5.1 – 6.0) | _ | <0.0001 |
|  | Mesh | 1,596 | 0.9 (0.8 – 1.0) | 82.7 (78.5, 86.1) |  |

Total fed= sum of fed alive, and fed dead captured for each month and combined for all the months; Mean= Average of fed mosquitoes caught per night per hut estimated as Williams mean due to skewness of mosquito count data; PE = Protective efficacy (1-IRR) *100 – it measures percentage reduction in mosquito successfully blood-feeding on a participant in the intervention relative to the control.
