## Supplemental Table 4 for "SC Johnson Guardian™ spatial repellent shows one-year efficacy against wild pyrethroid-resistant *Anopheles arabiensis*, with similar blood-feeding inhibition efficacy to Mosquito Shield™ in a Tanzanian experimental hut trial": Supplemental Table 4 .docx

**S4 Table** Monthly protective efficacy of Guardian in reducing the proportion of blood-fed wild pyrethroid-resistant *An. arabiensis*

| Month | Intervention | Total (n/N) | Mean (95% CI) | PE (95% CI) | p-value |
| --- | --- | --- | --- | --- | --- |
| 0 | Control | 186/1,294 | 12.1 (8.8 – 15.4) | _ | <0.001 |
|  | Guardian | 42/757 | 5.3 (2.8 – 7.8) | 67.5 (52.6, 77.7) |  |
| 1 | Control | 388/2,249 | 16.7 (13.4 – 20.0) |  | <0.001 |
|  | Guardian | 36/831 | 6.1 (3.1 – 9.1) | 79.1 (68.2, 86.2) |  |
| 2 | Control | 328/1,543 | 19.4 (15.0 – 23.8) |  | 0.0039 |
|  | Guardian | 29/248 | 7.3 (2.8 – 11.8) | 58.0 (24.3, 76.7) |  |
| 3 | Control | 280/1,545 | 25.1 (19.1 – 31.0) |  | <0.001 |
|  | Guardian | 29/447 | 7.8 (4.0 – 11.6) | 76.7 (64.4, 84.8) |  |
| 4 | Control | 830/2,918 | 27.0 (23.2 – 30.7) |  | <0.001 |
|  | Guardian | 78/930 | 9.5 (6.2 – 12.7) | 78.6 (67.2, 86.0) |  |
| 5 | Control | 562/1,780 | 30.7 (26.7 – 34.7) |  | <0.001 |
|  | Guardian | 51/609 | 9.6 (5.6 – 13.6) | 80.4 (67.6, 88.1) |  |
| 6 | Control | 331/707 | 44.4 (39.9 – 48.8) |  | <0.001 |
|  | Guardian | 49/302 | 15.8 (9.7 – 21.9) | 78.8 (66.9, 86.5) |  |
| 7 | Control | 432/920 | 46.8 (41.7 – 52.0) |  | <0.001 |
|  | Guardian | 63/425 | 15.7 (10.5 – 20.8) | 81.7 (71.9, 88.1) |  |
| 8 | Control | 677/1,538 | 45.5 (40.2 – 50.8) |  | <0.001 |
|  | Guardian | 91/749 | 14.8 (9.3 – 20.3) | 81.4 (75.9, 85.7) |  |
| 9 | Control | 1,066/4,718 | 25.1 (21.1 – 29.1) |  | <0.001 |
|  | Guardian | 358/2,826 | 15.7 (11.9 – 19.6) | 51.7 (35.0, 64.1) |  |
| 10 | Control | 812/2,807 | 28.8 (26.1 – 31.5) |  | 0.001 |
|  | Guardian | 299/1,930 | 17.7 (13.6 – 21.9) | 51.3 (39.6, 60.7) |  |
| 11 | Control | 1,002/3,127 | 32.2 (28.6 – 35.8) |  | <0.001 |
|  | Guardian | 335/1,930 | 16.4 (12.0 – 20.8) | 68.9 (51.3, 80.1) |  |
| 12 | Control | 403/1,784 | 29.0 (22.1 – 35.8) |  | 0.0027 |
|  | Guardian | 136/870 | 22.1 (14.8 – 29.4) | 43.4 (17.9, 61.0) |  |
| Overall | Control | 7,297/26,930 | 29.9 (28.4 – 31.4) |  | <0.001 |
|  | Guardian | 1,596/12,863 | 12.7 (11.4 – 14.1) | 67.0 (59.0, 73.0) |  |

n= sum of fed alive, and fed dead captured; N= overall total number of mosquitoes collected from huts (sum of fed alive, fed dead, unfed alive and unfed dead captured) for each month and combined for all the months; Mean= average proportion of fed mosquitoes caught per night per hut estimated as arithmetic mean for each month and combined for all the months; PE = Protective efficacy (1-OR) *100 – it measures percentage reduction in the odds of mosquitoes successfully blood-feeding on a participant in the intervention relative to the control.
