## Supplemental Table 5 for "SC Johnson Guardian™ spatial repellent shows one-year efficacy against wild pyrethroid-resistant *Anopheles arabiensis*, with similar blood-feeding inhibition efficacy to Mosquito Shield™ in a Tanzanian experimental hut trial": Supplemental Table 5 .docx

**S5 Table** Monthly mortality estimates of wild pyrethroid-resistant *An. arabiensis* induced by Guardian

| Month | Intervention | Total (n/N) | % 24-hour Mortality  (95% CI) | p-value |
| --- | --- | --- | --- | --- |
| 0 | Control | 21/1,294 | 3.0 (0.5 – 5.4) | <0.001 |
|  | Guardian | 151/757 | 15.0 (10.5 - 19.5) |  |
| 1 | Control | 7/2,249 | 0.3 (0.1 - 0.6) | <0.001 |
|  | Guardian | 187/831 | 22.0 (16.3 - 27.8) |  |
| 2 | Control | 2/1,543 | 0.1 (0.0 - 0.2) | <0.001 |
|  | Guardian | 44/248 | 12.9 (7.9 - 17.8) |  |
| 3 | Control | 1/1,545 | 0.1 (0.0 - 0.4) | <0.001 |
|  | Guardian | 26/447 | 5.8 (2.9 - 8.7) |  |
| 4 | Control | 20/2,918 | 0.5 (0.2 - 0.8) | <0.001 |
|  | Guardian | 289/930 | 29.2 (22.7 – 35.6) |  |
| 5 | Control | 3/1,780 | 0.2 (0.0 - 0.4) | <0.001 |
|  | Guardian | 257/609 | 35.5 (28.0 – 43.0) |  |
| 6 | Control | 2/707 | 0.6 (0.0 – 1.9) | <0.001 |
|  | Guardian | 118/302 | 30.6 (23.0 – 38.2) |  |
| 7 | Control | 1/920 | 0.0 (0.0 - 0.1) | <0.001 |
|  | Guardian | 170/425 | 32.3 (24.7 – 40.0) |  |
| 8 | Control | 4/1,538 | 0.5 (0.0 – 1.3) | <0.001 |
|  | Guardian | 242/749 | 29.4 (23.1 – 35.7) |  |
| 9 | Control | 3/4,718 | 0.1 (0.0 – 0.1) | <0.001 |
|  | Guardian | 283/2,826 | 15.9 (10.9 – 20.8) |  |
| 10 | Control | 4/2,807 | 0.3 (0.0 – 0.7) | <0.001 |
|  | Guardian | 204/1,930 | 13.2 (9.9 – 16.6) |  |
| 11 | Control | 4/3,127 | 0.1 (0.0 – 0.2) | <0.001 |
|  | Guardian | 128/1,930 | 9.2 (6.6 – 11.9) |  |
| 12 | Control | 2/1,784 | 0.1 (0.0 – 0.2) | <0.001 |
|  | Guardian | 71/870 | 9.0 (4.8 – 13.2) |  |
| Overall | Control | 74/26,930 | 0.5 (0.2 – 0.7) | <0.001 |
|  | Guardian | 2,170/12,863 | 20.1 (18.4 – 21.8) |  |

n= total number of mosquitoes that died over 24-hours holding period including dead fed and dead unfed collected the morning after from the huts; N= Overall total number of mosquitoes collected from huts (sum of fed alive, fed dead, unfed alive and unfed dead captured); p-value from logistic regression comparing odds of a mosquito dying in the huts with a treatment relative to those without.
