## Supplemental Figure 1 for "SC Johnson Guardian™ spatial repellent shows one-year efficacy against wild pyrethroid-resistant *Anopheles arabiensis*, with similar blood-feeding inhibition efficacy to Mosquito Shield™ in a Tanzanian experimental hut trial"

| | 2022 | | | | | | | | 2023 | | | | |
|---|---|---|---|---|---|---|---|---|---|---|---|---|---|
| | May | Jun | Jul | Aug | Sep | Oct | Nov | Dec | Jan | Feb | Mar | Apr | May |
| Guardian | | | | | | | | | | | | | |
| Mosquito Shield | | | | | | | | | | | | | |
